# Cardiac Measurement Calculation on Point-of-Care Ultrasonography with Artificial Intelligence

**DOI:** 10.1101/2025.06.27.25330438

**Authors:** Sarah F Mercaldo, Bernardo C Bizzo, Tsion Sadore, Madeleine A Halle, Ashley L MacDonald, Isabella Newbury-Chaet, Eric L’Italien, Alex S Schultz, Victor Tam, Sheila M Hegde, Judy R Mangion, Praveen Mehrotra, Qiong Zhao, Justina Wu, James M Hillis

**Affiliations:** Digital Clinical Research Organization, Mass General Brigham AI, Boston, MA, USA; Department of Radiology, Massachusetts General Hospital, Boston, MA, USA; Harvard Medical School, Boston, MA, USA; Division of Cardiovascular Medicine, Department of Medicine, Brigham and Women’s Hospital, Boston, MA, USA; Division of Cardiology, Sidney Kimmel Medical College, Thomas Jefferson University, PA; Inova Schar Heart and Vascular, Inova Health System, VA; Department of Neurology, Massachusetts General Hospital, Boston, MA, USA

## Abstract

**Introduction:** Point-of-care ultrasonography (POCUS) enables clinicians to obtain critical diagnostic information at the bedside especially in resource limited settings. This information may include 2D cardiac quantitative data, although measuring the data manually can be time-consuming and subject to user experience. Artificial intelligence (AI) can potentially automate this quantification. This study assessed the interpretation of key cardiac measurements on POCUS images by an AI-enabled device (AISAP Cardio V1.0).

**Methods:** This retrospective diagnostic accuracy study included 200 POCUS cases from four hospitals (two in Israel and two in the United States). Each case was independently interpreted by three cardiologists and the device for seven measurements (left ventricular (LV) ejection fraction, inferior vena cava (IVC) maximal diameter, left atrial (LA) area, right atrial (RA) area, LV end diastolic diameter, right ventricular (RV) fractional area change and aortic root diameter). The endpoints were the root mean square error (RMSE) of the device compared to the average cardiologist measurement (LV ejection fraction and IVC maximal diameter were primary endpoints; the other measurements were secondary endpoints). Predefined passing criteria were based on the upper bounds of the RMSE 95% confidence intervals (CIs). The inter-cardiologist RMSE was also calculated for reference.

**Results:** The device achieved the passing criteria for six of the seven measurements. While not achieving the passing criterion for RV fractional area change, it still achieved a better RMSE than the inter-cardiologist RMSE. The RMSE was 6.20% (95% CI: 5.57 to 6.83; inter-cardiologist RMSE of 8.23%) for LV ejection fraction, 0.25cm (95% CI: 0.20 to 0.29; 0.36cm) for IVC maximal diameter, 2.39cm^2^ (95% CI: 1.96 to 2.82; 4.39cm^2^) for LA area, 2.11cm^2^ (95% CI: 1.75 to 2.47; 3.49cm^2^) for RA area, 5.06mm (95% CI: 4.58 to 5.55; 4.67mm) for LV end diastolic diameter, 10.17% (95% CI: 9.01 to 11.33; 14.12%) for RV fractional area change and 0.19cm (95% CI: 0.16 to 0.21; 0.24cm) for aortic root diameter.

**Discussion:** The device accurately calculated these cardiac measurements especially when benchmarked against inter-cardiologist variability. Its use could assist clinicians who utilize POCUS and better enable their clinical decision-making.

## Introduction

Point-of-care ultrasonography (POCUS) has revolutionized many aspects of acute clinical care over recent decades.^1–3^ It enables clinicians to obtain more diagnostic information at the bedside, and has been adopted by multiple specialties especially emergency medicine and hospital internal medicine.^4–6^ With some handheld probes connecting directly to a smartphone or tablet, it can be particularly beneficial in resource limited settings.^7–10^ Its use includes the quantification of key cardiac measurements such as left ventricular (LV) ejection fraction and inferior vena cava (IVC) size.^11^

Artificial intelligence (AI) has similarly started to revolutionize clinical care with many of its current applications relating to medical image interpretation.^12,13^ One of its core uses is the automatic quantification of measurements, especially those that otherwise require time-consuming manual tracings. It has previously been used to quantify cardiac measurements using conventional echocardiograms.^14–16^ It has also been shown to improve interobserver variability and workflow efficiency for echocardiogram interpretation.^17,18^ Echocardiography, however, typically provides more comprehensive evaluations with a higher image resolution than POCUS.^19^

This study evaluated the ability of an AI-enabled device to calculate seven key cardiac measurements based on POCUS images including LV ejection fraction, IVC maximal diameter, left atrial (LA) area, right atrial (RA) area, LV end diastolic diameter, right ventricular (RV) fractional area change and aortic root diameter.

## Methods

### Study design

This diagnostic study was conducted using POCUS cases acquired prospectively from four hospitals in Israel and the United States. The acquisition of cases was approved by local Institutional Review Boards at each of Baruch Padeh Medical Center, Inova Health, Sheba Medical Center and Thomas Jefferson University Hospital. All patients provided informed consent. The subsequent diagnostic study involved transfer of deidentified cases to Mass General Brigham and was approved by the Mass General Brigham Institutional Review Board. The study was conducted in accordance with relevant guidelines and regulations. This manuscript followed the Standards for Reporting of Diagnostic Accuracy (STARD) reporting guideline.

### Inclusion criteria

Eligible participants were identified in a consecutive manner in both the inpatient and outpatient settings. They were patients with suspected or established cardiac pathology, and with an indication for cardiac ultrasound. They were initially identified by treating physicians including cardiologists, internal medicine physicians and emergency department physicians, and were consented by local research personnel. The inclusion criteria included male or female individuals aged 21 or older, the provision of a signed and dated informed consent form, a stated willingness to comply with all study procedures and availability for the study duration. The exclusion criteria included unstable clinical condition as determined by the principal investigator (where ultrasonography was considered detrimental to the participant), clinical conditions that prevented an ultrasonography study according to the investigators or treating physicians, presence of a prosthetic cardiac valve of any type or position, pregnancy or lactation, contact isolation due to infectious disease, presence of a left ventricular assist device, known congenital heart disease, body mass index (BMI) of 40 or greater, history of lung resection, known very poor cardiac ultrasonography image quality based on prior comprehensive echocardiography examinations, or mechanical ventilation or other situations where the required ultrasonography views could not be adequately obtained.

### POCUS

The POCUS cases were acquired by physicians and sonographers at the bedside using mostly handheld devices. They contained only four cardiac views according to the American Society of Echocardiography guidelines.^21^ These views included parasternal long axis with and without color Doppler, apical four chamber with and without color Doppler, parasternal short axis at the aortic valve level, and subcostal IVC view.

### Cohort selection

Once cases were acquired at individual sites, they underwent de-identification prior to transfer to the sponsor. These cases were maintained separately from the cases used for device development. Only two of the four sites had contributed cases for device development, which had all been acquired prior to the cases used for the validation study. The deidentified cases were subsequently transferred to Mass General Brigham for the diagnostic study. The final cohort required only a subset of cases based on powering calculations, which was selected in a randomized manner.

### Ground truth process

The three ground truth cardiologists were US board-certified with a minimum of 5 years of experience in echocardiography. They received training for this project and performed their annotations using a validated custom interface in Siemens syngo.via (version VA20). They reviewed the available loops of the relevant view(s) for each measurement (Supplementary Table 1) and chose the most adequate loop to perform the measurement. They did not receive additional clinical information about the cases. For LV ejection fraction, they visually estimated the measurement; other measurements were calculated based on the American Society of Echocardiography guidelines.^21^ RV fractional area change was calculated from manually traced end-diastolic and end-systolic RV areas. The component measurements for RV fractional area change were measured 3 times on different beats and the average fractional area change was calculated; each cardiologist obtained the other measurements once per case.

Separately they indicated when a measurement could not be obtained and should therefore be excluded; if at least two of the cardiologists indicated that the measurement could not be obtained then the measurement was excluded for that case; if only one of them indicated that the measurement could be obtained then that cardiologist was asked to review the case and make an actual best determination of the measurement.

### Device development and inference

The device evaluated in this study was trained using labeled echocardiography data acquired at Baruch Padeh Medical Center and Sheba Medical Center between 2009 and 2020. The development set included 145,217 adult patient examinations, which were split into a training set (95,167 cases, 65.5%), validation set (29,090 cases, 20.0%), and a hold-out test set (20,960 cases, 14.4%; which was separate to the cases evaluated in the current study). This development set represented a mix of inpatient and outpatient studies on a range of echocardiography machines. Each examination comprised 20-80 ultrasound images and loops, capturing various cardiac views according to echocardiography protocols.

In brief, the device architecture consists of 2 types of models. The first is a model which has a combination of convolutional neural networks (CNNs) and transformers. Feature vectors are extracted using designated trained EfficientNetV2^22^ like architecture and fed into transformer blocks with connected CNNs to highlight relevant features across time. The final output vectors then underwent max-pooling and were processed by a regressor head. The second type of model is an instance segmentation CNN model trained for segmentation of relevant anatomical parts within an ultrasound image.

The device analyzed parasternal, apical and subcostal views; for measurements requiring multiple views, frames from both views were concatenated. The presence of multiple loops of a view during inference resulted in averaged predictions in an effort to increase the robustness of measurements. The device performed inference independently of the ground truth cardiologists and did not receive additional clinical information about the cases.

The device provided a single value for each of the 7 measurements. It could also provide an output of “NULL” that reflected it could not interpret that measurement for the case. The reasons included insufficient quality of images (as the device sees them) or inability to calculate the measurement due to the absence of key points in the image used by the device.

### Statistical Analysis

The statistical analysis was conducted using software R version 4.2.0 (R Foundation for Statistical Computing, Vienna, Austria, 2019). The predefined primary and secondary endpoints were the diagnostic accuracy of the device as determined by the root mean square error (RMSE) when compared with the average of the three ground truth cardiologists. LV ejection fraction and IVC maximal diameter were considered primary endpoints given these metrics often impact patient care more directly; the other measurements were considered secondary endpoints. The RMSE was calculated as the square root of the mean of the squared differences between the device measurements and the average cardiologist measurements, providing a measure of overall deviation.^23^ The inter-cardiologist RMSE was also calculated for reference, which was the average of the pairwise RMSEs between each combination of cardiologists. The passing criteria for the primary endpoints were RMSEs of less than 7.0% for LV ejection fraction and 0.3cm for IVC maximal diameter, which were to be met by the upper bounds of the 95% confidence intervals (CIs). The passing criteria for the secondary endpoints were RMSEs less than 20% of the average of the absolute measurement from the three cardiologists, which similarly was to be met by the upper bounds of the 95% CIs.

To further evaluate the interchangeability and accuracy of the device measurements compared to cardiologist measurements, additional predefined secondary assessments were calculated including the individual equivalence coefficient (IEC; to compare variability of the relative differences between the device and the cardiologists with the variability of the relative differences between cardiologists),^24^ mean absolute error (MAE; to assess the average magnitude of errors),^23^ limits of agreement (LOA) using Bland-Altman analysis (to evaluate the presence of systematic errors between the device and cardiologists),^25^ and intraclass correlation coefficient (ICC; to measure the consistency and reproducibility of the measurements).^26^ MAE was computed as the mean of the absolute differences between the device and cardiologist measurements, providing a metric of average error magnitude without being influenced by the direction of errors. The Bland-Altman analysis involved plotting the mean of each device-cardiologist measurement pair against their difference, allowing for visual assessment of bias and agreement. The 95% LOAs were defined as the mean difference ± 1.96 × standard deviation of the differences, indicating the range within which most differences are expected to lie. For the IEC and ICC calculations, the measurements from each of the cardiologists were considered. For MAE and LOA, the device was compared with the average measurement from the three cardiologists.

Subgroup analyses were calculated for demographic (sex, age, race and ethnicity, BMI), disease (LV ejection fraction) and image (ultrasound probe manufacturer, image quality score, role of person acquiring scan) parameters. The subgroups were predefined except for race and role of person acquiring scan (which were added as post-hoc analyses), and BMI and LV ejection fraction (for which the subgroups were updated to have more subgroups following regulatory feedback). Each ultrasound probe manufacturer only had a single model so both the manufacturers and models are specified in the results.

All metrics were accompanied by bootstrapped 95% CIs with 2,000 resamples. An emphasis was placed on the interpretation of CIs to infer the reliability of calculations. No adjustments for multiple comparisons were made. Sample size calculations were performed a priori for the primary endpoints and powered based on preliminary device results.

## Results

### Participant characteristics

The overall cohort included cases from 200 patients (Figure 1). The mean age (± standard deviation) was 65.4 (± 16.0) years and 74 (37.0%) were female patients (Table 1). A case was excluded from the analysis for a given measurement if the cardiologists or the device could not obtain the measurement (Supplementary Table 2).

**Figure 1:**
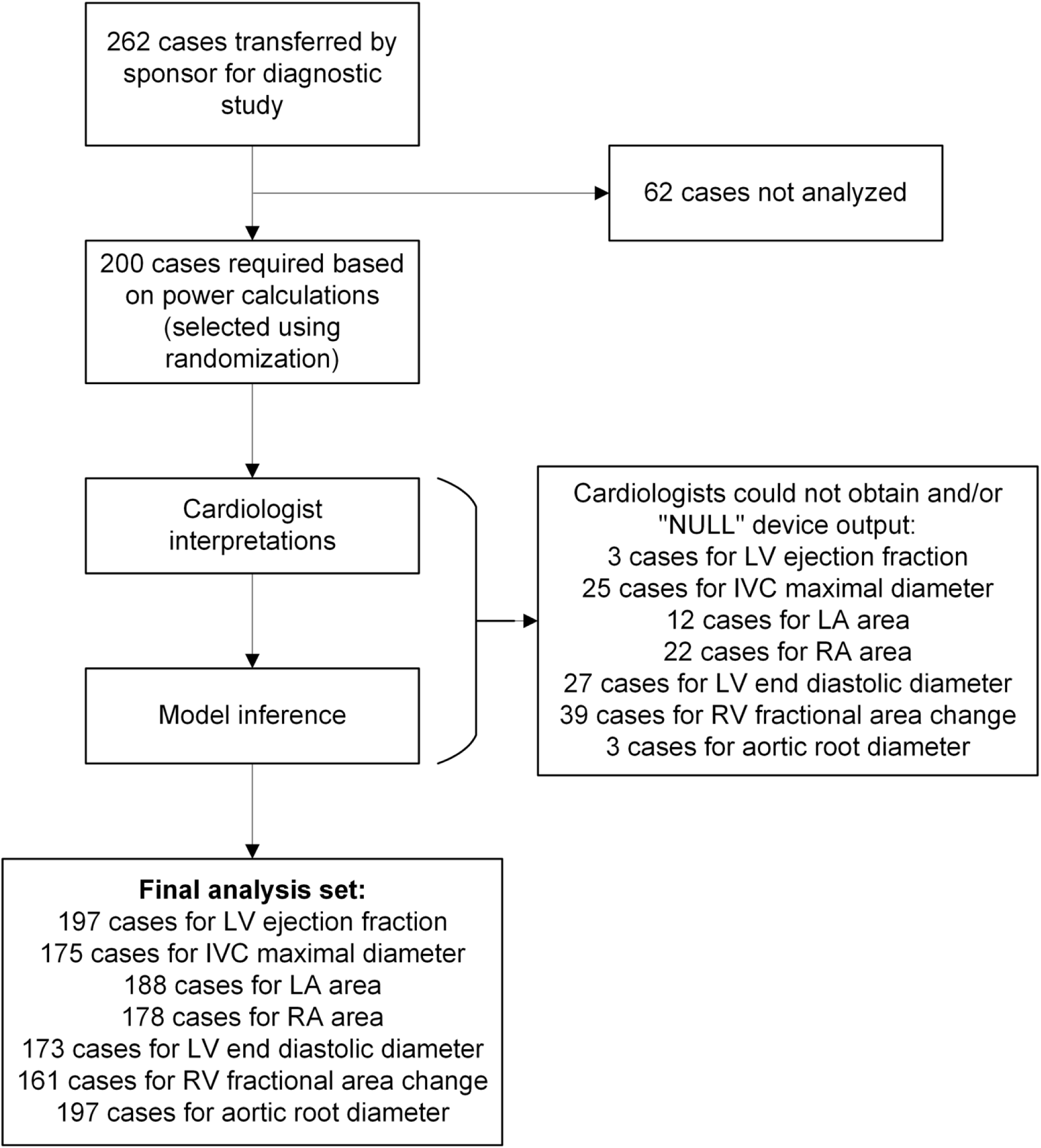
Cohort selection. Abbreviations: IVC, inferior vena cava; LA, left atrial; LV, left ventricular; RA, right atrial; RV, right ventricular.

**Table 1:** Baseline demographic and technical information. Abbreviations: BMI, body mass index; LV, left ventricular; SD, standard deviation.

|  | Number of cases (%) |
| --- | --- |
| **Total cohort** | 200 |
| **Age** |  |
| ≤65 years | 84 (42.0%) |
| >65 years | 116 (58.0%) |
| Mean (± SD) | 65.4 (± 16.0) |
| **Sex** |  |
| Female | 74 (37.0%) |
| Male | 126 (63.0%) |
| **Race and Ethnicity** |  |
| American Indian or Alaska Native | 2 (1.0%) |
| Asian | 11 (5.5%) |
| Black or African American | 26 (13.0%) |
| Caucasian | 151 (75.5%) |
| Hispanic or Latino | 8 (4.0%) |
| Mixed | 1 (0.5%) |
| Other | 1 (0.5%) |
| **BMI** |  |
| <25 | 68 (34.0%) |
| 25 to <30 | 82 (41.0%) |
| 30 to <35 | 37 (18.5%) |
| ≥35 | 11 (5.5%) |
| Unknown | 2 (1.0%) |
| Mean (± SD) | 27.2 (± 4.7) |
| **LV ejection fraction** |  |
| <30% | 19 (9.6%) |
| ≥30% to <53% | 49 (24.7%) |
| ≥53% | 130 (65.7%) |
| **Ultrasound Probe Manufacturer (Model)** |  |
| EchoNous (Kosmos) | 37 (18.5%) |
| Philips (Lumify) | 82 (41.0%) |
| Wisonic (Navi) | 25 (12.5%) |
| GE Healthcare (Venue) | 56 (28.0%) |
| **Image quality score** |  |
| 1 | 1 (0.5%) |
| 2 | 2 (1.0%) |
| 3 | 11 (5.5%) |
| 4 | 36 (18.0%) |
| 5 | 99 (49.5%) |
| Unknown | 51 (25.5%) |
| **Role of person acquiring scan** |  |
| Physician | 37 (18.5%) |
| Sonographer | 183 (81.5%) |

### Primary and secondary endpoint analysis

The RMSE was calculated between the device and the three cardiologists (Table 2). Both primary endpoint passing criteria were met, with the RMSEs for LV ejection fraction and IVC maximal diameter having a 95% CI upper bound less than 7.0% and 0.3cm respectively. Four out of the five secondary endpoint passing criteria were met, with the RMSEs achieving a 95% CI upper bound less than 20% of the mean cardiologist measurement. RV fractional area change was the one secondary endpoint that did not achieve its passing criterion, although the RMSE was still better between the device and cardiologists than between the cardiologists. The other measurements, except for LV end diastolic diameter, also had better RMSEs between the device and the cardiologists than between the cardiologists.

**Table 2:**
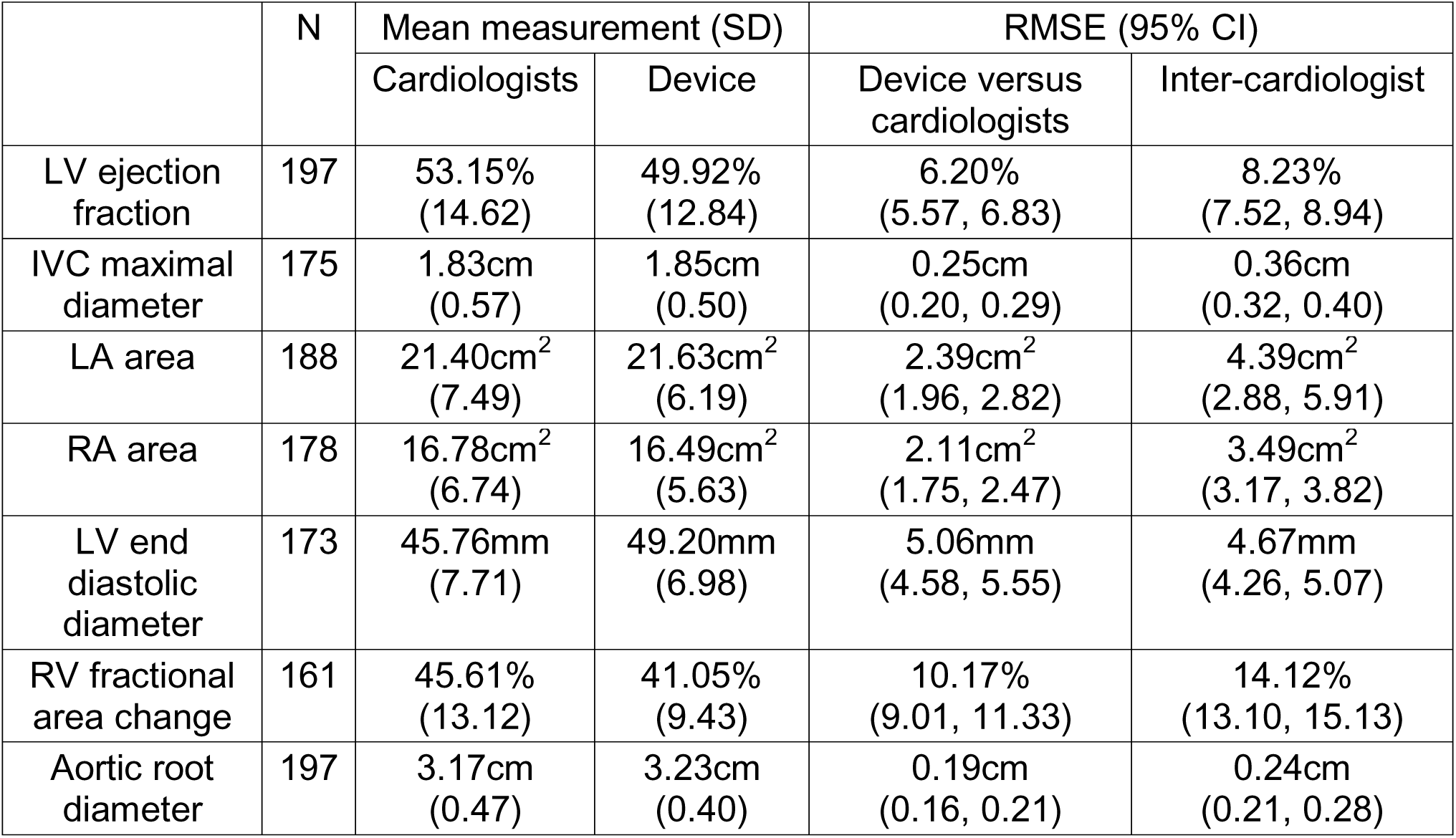
Primary and secondary endpoint results. The RMSE between the device and the average of the three cardiologists for the seven measurements (LV ejection fraction and IVC maximal diameter were primary endpoints; the others were secondary endpoints). The inter-cardiologist RMSE is provided for reference. This table also incorporates the number of cases and mean measurements for each measurement. Abbreviations: CI, confidence interval; IVC, inferior vena cava; LA, left atrial; LV, left ventricular; N, number of cases; RA, right atrial; RMSE, root mean square error; RV, right ventricular; SD, standard deviation.

### Secondary assessment analysis

The IEC, MAE, LOA and ICC were calculated between the device and the three cardiologists (Table 3). The IECs were mostly less than 0, showing better variability of the relative differences between the device and cardiologists compared with the variability of the relative differences between the cardiologists. The MAEs, which provided an additional statistical method to RMSEs for measuring the error of the device, were less than the RMSEs. The LOAs, which can show the presence of systematic bias in a device, had mean differences that were unlikely to be clinically meaningful. The ICCs were mostly above 0.75, reflecting the strength of the correlation between the measurements from the device and the measurements from the cardiologists.

**Table 3:** Secondary assessment results. The IEC, MAE, LOA and ICC between the device and the three cardiologists for the seven measurements. The LOA is expressed as the mean difference ± 1.96 × standard deviation of the differences. This table also incorporates the number of cases for each measurement. Abbreviations: CI, confidence interval; ICC, intraclass coefficient; IEC, individual equivalence coefficient; IVC, inferior vena cava; LA, left atrial; LOA, limits of agreement; LV, left ventricular; MAE, mean absolute error; N, number of cases; RA, right atrial; RV, right ventricular.

|  | N | IEC<br>(95% CI) | MAE<br>(95% CI) | LOA | ICC<br>(95% CI) |
| --- | --- | --- | --- | --- | --- |
| LV ejection fraction | 197 | -0.20<br>(-0.49, 0.09) | 5.01%<br>(4.48, 5.53) | 3.18 ±<br>10.47% | 0.84<br>(0.81, 0.88) |
| IVC maximal diameter | 175 | -0.42<br>(-0.82, -0.03) | 0.17cm<br>(0.14, 0.20) | -0.03 ±<br>0.48cm | 0.78<br>(0.72, 0.84) |
| LA area | 188 | -0.74<br>(-1.05, -0.43) | 1.67cm<sup>2</sup><br>(1.42, 1.91) | -0.39 ±<br>4.64cm<sup>2</sup> | 0.84<br>(0.75, 0.93) |
| RA area | 178 | -0.69<br>(-0.99, -0.40) | 1.48cm<sup>2</sup><br>(1.27, 1.70) | -0.03 ±<br>4.15 cm<sup>2</sup> | 0.85<br>(0.80, 0.90) |
| LV end diastolic diameter | 173 | 0.93<br>(0.35, 1.51) | 4.09mm<br>(3.65, 4.54) | -3.46 ±<br>7.28mm | 0.77<br>(0.73, 0.82) |
| RV fractional area change | 161 | -0.33<br>(-0.59, -0.06) | 7.91%<br>(6.93, 8.89) | 4.77 ±<br>17.67% | 0.42<br>(0.33, 0.51) |
| Aortic root diameter | 197 | -0.16<br>(-0.61, 0.30) | 0.14cm<br>(0.13, 0.16) | -0.07 ±<br>0.34cm | 0.86<br>(0.82, 0.90) |

### Subgroup analyses

The subgroup analyses evaluated the RMSEs across demographic, disease and image parameter subgroups (Table 4). The vast majority of RMSEs were similar across subgroups as suggested by overlapping ranges of 95% CIs: there were overlapping ranges for all measurements in the subgroups for age, sex, race and ethnicity, image quality score and role of person acquiring scan. The measurements with non-overlapping ranges included RA area for BMI subgroups (the <25 subgroup had lower RMSE than the [30,35) subgroup), LV ejection fraction for LV ejection fraction subgroups (the <30% subgroup had lower RMSE than the ≥30% to <53% and ≥53% subgroups), LV end diastolic diameter for LV ejection fraction subgroups (the <30% subgroup had lower RMSE than the ≥53% subgroup), and LV ejection fraction for manufacturer subgroups (the GE Healthcare subgroup had lower RMSE than the Philips or Wisonic subgroups). In addition, the BMI unknown subgroup had only 2 cases and often had non-overlapping 95% CIs.

**Table 4:** Subgroup analysis results. The RMSE between the device and the average of the three cardiologists for the seven measurements for demographic, disease and image parameter subgroups. Abbreviations: BMI, body mass index; CI, confidence interval; IVC, inferior vena cava; LA, left atrial; LV, left ventricular; N, number of cases; RA, right atrial; RMSE, root mean square error; RV, right ventricular.

|  | RMSE<br>(95% CI; N) |  |  |  |  |  |  |
| --- | --- | --- | --- | --- | --- | --- | --- |
|  | LV ejection fraction | IVC maximal diameter | LA area | RA area | LV end diastolic diameter | RV fractional area change | Aortic root diameter |
| Age |  |  |  |  |  |  |  |
| ≤65 years | 5.61%<br>(4.50, 6.73;<br>82) | 0.26cm<br>(0.17, 0.34;<br>74) | 2.35cm<sup>2</sup><br>(1.42, 3.29;<br>79) | 2.13cm<sup>2</sup><br>(1.50, 2.76;<br>78) | 4.76mm<br>(4.04, 5.48;<br>77) | 11.03%<br>(9.22, 12.85;<br>67) | 0.17cm<br>(0.12, 0.21;<br>84) |
| >65 years | 6.59%<br>(5.86, 7.32;<br>115) | 0.24cm<br>(0.20, 0.28;<br>101) | 2.42cm<sup>2</sup><br>(2.05, 2.79;<br>109) | 2.10cm<sup>2</sup><br>(1.68, 2.52;<br>100) | 5.29mm<br>(4.64, 5.95;<br>96) | 9.51%<br>(8.03, 11.00;<br>94) | 0.20cm<br>(0.17, 0.23;<br>113) |
| Sex |  |  |  |  |  |  |  |
| Female | 6.45%<br>(5.44, 7.46;<br>73) | 0.23cm<br>(0.17, 0.28;<br>66) | 2.46cm<sup>2</sup><br>(1.61, 3.31;<br>69) | 1.78cm<sup>2</sup><br>(1.01, 2.54;<br>66) | 4.60mm<br>(3.79, 5.40;<br>65) | 9.29%<br>(7.71, 10.86;<br>59) | 0.19cm<br>(0.16, 0.22;<br>72) |
| Male | 6.05%<br>(5.26, 6.84;<br>124) | 0.26cm<br>(0.20, 0.32;<br>109) | 2.35cm<sup>2</sup><br>(1.91, 2.79;<br>119) | 2.29cm<sup>2</sup><br>(1.88, 2.69;<br>112) | 5.33mm<br>(4.73, 5.92;<br>108) | 10.65%<br>(9.06, 12.25;<br>102) | 0.19cm<br>(0.15, 0.22;<br>125) |
| Race and Ethnicity |  |  |  |  |  |  |  |
| Black or African American | 7.77%<br>(5.24, 10.30;<br>25) | 0.20cm<br>(0.08, 0.32;<br>22) | 2.05cm<sup>2</sup><br>(1.49, 2.62;<br>23) | 2.19cm<sup>2</sup><br>(1.17, 3.21;<br>22) | 4.37mm<br>(2.95, 5.78;<br>25) | 9.71%<br>(7.43, 11.98;<br>19) | 0.20cm<br>(0.15, 0.25;<br>26) |
| Caucasian | 6.14%<br>(5.54, 6.74;<br>150) | 0.25cm<br>(0.20, 0.31;<br>132) | 2.36cm<sup>2</sup><br>(1.86, 2.87;<br>145) | 2.07cm<sup>2</sup><br>(1.65, 2.50;<br>135) | 5.34mm<br>(4.80, 5.87;<br>126) | 9.64%<br>(8.35, 10.93;<br>124) | 0.19cm<br>(0.16, 0.22;<br>148) |
| Other races (combined) | 4.38%<br>(3.11, 5.65;<br>22) | 0.24cm<br>(0.15, 0.33;<br>21) | 2.90cm<sup>2</sup><br>(1.50, 4.30;<br>20) | 2.28cm<sup>2</sup><br>(1.13, 3.44;<br>21) | 4.10mm<br>(2.53, 5.67;<br>22) | 13.66%<br>(9.33, 17.98;<br>18) | 0.17cm<br>(0.12, 0.23;<br>23) |
| BMI |  |  |  |  |  |  |  |
| <25 | 6.49%<br>(5.42, 7.57;<br>67) | 0.21cm<br>(0.16, 0.27;<br>60) | 2.32cm<sup>2</sup><br>(1.78, 2.86;<br>64) | 1.43cm<sup>2</sup><br>(1.09, 1.77;<br>58) | 4.56mm<br>(3.84, 5.28;<br>58) | 9.15%<br>(7.02, 11.28;<br>57) | 0.19cm<br>(0.15, 0.22;<br>67) |
| [25,30) | 6.11%<br>(5.05, 7.17;<br>81) | 0.28cm<br>(0.20, 0.36;<br>75) | 2.62cm<sup>2</sup><br>(1.79, 3.44;<br>78) | 1.92cm<sup>2</sup><br>(1.47, 2.37;<br>74) | 5.33mm<br>(4.58, 6.07;<br>73) | 11.39%<br>(9.58, 13.20;<br>68) | 0.15cm<br>(0.12, 0.18;<br>81) |
| [30,35) | 6.29%<br>(5.00, 7.57;<br>36) | 0.24cm<br>(0.17, 0.30;<br>30) | 2.12cm<sup>2</sup><br>(1.61, 2.63;<br>33) | 3.03cm<sup>2</sup><br>(2.05, 4.02;<br>34) | 5.22mm<br>(3.92, 6.52;<br>29) | 9.37%<br>(6.96, 11.78;<br>27) | 0.24cm<br>(0.17, 0.31;<br>36) |
| ≥35 | 5.01%<br>(3.65, 6.37;<br>11) | 0.16cm<br>(0.05, 0.27;<br>8) | 2.10cm<sup>2</sup><br>(1.17, 3.04;<br>11) | 2.92cm<sup>2</sup><br>(1.21, 4.64;<br>10) | 5.59mm<br>(3.44, 7.74;<br>11) | 8.60%<br>(3.03, 14.18;<br>7) | 0.23cm<br>(0.14, 0.32;<br>11) |
| Unknown | 4.16% | 0.24cm | 0.43cm<sup>2</sup> | 1.18cm<sup>2</sup> | 3.51mm | 9.53% | 0.03cm |
|  | (4.13, 4.20;<br>2) | (0.00, 0.47;<br>2) | (0.08, 0.79;<br>2) | (0.55, 1.82;<br>2) | (2.13, 4.89;<br>2) | (2.25, 16.81;<br>2) | (0.03, 0.03;<br>2) |
| LV ejection fraction |  |  |  |  |  |  |  |
| <30% | 4.29%<br>(3.24, 5.33;<br>19) | 0.27cm<br>(0.17, 0.38;<br>16) | 3.45cm<sup>2</sup><br>(1.83, 5.06;<br>18) | 1.78cm<sup>2</sup><br>(1.25, 2.31;<br>17) | 3.57mm<br>(2.28, 4.85;<br>14) | 10.74%<br>(7.42, 14.06;<br>14) | 0.14cm<br>(0.08, 0.19;<br>19) |
| ≥30% to<br><53% | 7.04%<br>(5.79, 8.29;<br>49) | 0.18cm<br>(0.14, 0.22;<br>41) | 1.63cm<sup>2</sup><br>(1.26, 2.00;<br>46) | 1.79cm<sup>2</sup><br>(1.11, 2.47;<br>45) | 4.44mm<br>(3.53, 5.34;<br>46) | 9.75%<br>(7.35, 12.15;<br>38) | 0.19cm<br>(0.14, 0.24;<br>49) |
| ≥53% | 6.10%<br>(5.33, 6.87;<br>129) | 0.26cm<br>(0.20, 0.32;<br>117) | 2.45cm<sup>2</sup><br>(1.92, 2.98;<br>123) | 2.27cm<sup>2</sup><br>(1.79, 2.75;<br>115) | 5.46mm<br>(4.85, 6.07;<br>111) | 10.24%<br>(8.78, 11.71;<br>109) | 0.19cm<br>(0.16, 0.22;<br>127) |
| Ultrasound Probe Manufacturer (Model) |  |  |  |  |  |  |  |
| EchoNous (Kosmos) | 5.20%<br>(4.20, 6.21;<br>37) | 0.30cm<br>(0.15, 0.45;<br>34) | 2.54cm<sup>2</sup><br>(0.99, 4.08;<br>36) | 1.76cm<sup>2</sup><br>(1.40, 2.13;<br>34) | 5.18mm<br>(4.02, 6.35;<br>32) | 8.50%<br>(6.29, 10.71;<br>25) | 0.15cm<br>(0.12, 0.19;<br>37) |
| Philips (Lumify) | 7.12%<br>(6.01, 8.23;<br>80) | 0.23cm<br>(0.17, 0.28;<br>70) | 1.96cm<sup>2</sup><br>(1.50, 2.43;<br>74) | 1.86cm<sup>2</sup><br>(1.42, 2.31;<br>71) | 5.10mm<br>(4.32, 5.88;<br>70) | 9.71%<br>(8.27, 11.15;<br>66) | 0.21cm<br>(0.16, 0.25;<br>80) |
| Wisonic (Navi) | 7.28%<br>(5.93, 8.63;<br>25) | 0.19cm<br>(0.13, 0.25;<br>23) | 2.62cm<sup>2</sup><br>(2.00, 3.24;<br>25) | 1.90cm<sup>2</sup><br>(1.22, 2.59;<br>24) | 5.09mm<br>(3.54, 6.64;<br>18) | 8.77%<br>(5.72, 11.82;<br>22) | 0.18cm<br>(0.13, 0.23;<br>25) |
| GE Healthcare (Venue) | 4.67%<br>(3.75, 5.59;<br>55) | 0.25cm<br>(0.19, 0.31;<br>48) | 2.70cm<sup>2</sup><br>(1.97, 3.43;<br>53) | 2.69cm<sup>2</sup><br>(1.78, 3.60;<br>49) | 4.94mm<br>(4.11, 5.76;<br>53) | 12.03%<br>(9.51, 14.54;<br>48) | 0.18cm<br>(0.14, 0.22;<br>55) |
| Image quality score |  |  |  |  |  |  |  |
| ≤4 | 5.68%<br>(4.49, 6.87;<br>48) | 0.26cm<br>(0.18, 0.33;<br>41) | 2.32cm<sup>2</sup><br>(1.79, 2.85;<br>45) | 2.27cm<sup>2</sup><br>(1.76, 2.78;<br>41) | 4.51mm<br>(3.83, 5.18;<br>44) | 9.07%<br>(6.56, 11.58;<br>39) | 0.19cm<br>(0.15, 0.23;<br>48) |
| >4 | 5.70%<br>(5.00, 6.40;<br>99) | 0.27cm<br>(0.19, 0.34;<br>89) | 2.62cm<sup>2</sup><br>(1.90, 3.35;<br>94) | 2.15cm<sup>2</sup><br>(1.53, 2.76;<br>92) | 5.02mm<br>(4.31, 5.73;<br>86) | 10.88%<br>(9.14, 12.63;<br>82) | 0.18cm<br>(0.14, 0.22;<br>98) |
| Unknown | 7.51%<br>(6.02, 8.99;<br>50) | 0.19cm<br>(0.14, 0.23;<br>45) | 1.95cm<sup>2</sup><br>(1.51, 2.39;<br>49) | 1.88cm<sup>2</sup><br>(1.41, 2.35;<br>45) | 5.66mm<br>(4.59, 6.73;<br>43) | 9.68%<br>(7.90, 11.46;<br>40) | 0.20cm<br>(0.16, 0.23;<br>51) |
| Role of person acquiring scan |  |  |  |  |  |  |  |
| Physician | 5.90%<br>(4.77, 7.03;<br>37) | 0.19cm<br>(0.14, 0.23;<br>35) | 1.83cm<sup>2</sup><br>(1.38, 2.28;<br>36) | 1.79cm<sup>2</sup><br>(1.33, 2.24;<br>36) | 5.12mm<br>(3.91, 6.33;<br>29) | 9.48%<br>(6.93, 12.03;<br>29) | 0.20cm<br>(0.12, 0.28;<br>36) |
| Sonographer | 6.27%<br>(5.58, 6.97;<br>160) | 0.26cm<br>(0.20, 0.31;<br>140) | 2.51cm<sup>2</sup><br>(2.02, 3.00;<br>152) | 2.19cm<sup>2</sup><br>(1.74, 2.63;<br>142) | 5.05mm<br>(4.51, 5.60;<br>144) | 10.32%<br>(9.03, 11.61;<br>132) | 0.18cm<br>(0.16, 0.21;<br>161) |

## Discussion

This study assessed the performance of an AI-enabled device at interpreting seven key cardiac measurements on images acquired using POCUS devices. It achieved both of its primary endpoint passing criteria and all but one of its secondary endpoint passing criteria, which were based on the RMSE between the device and the average of three cardiologists for the measurements. It also demonstrated better performance compared to the inter-cardiologist RMSE for all but one of the measurements. Importantly, this study was based mostly on images acquired from inexpensive, predominantly handheld devices that could perform a short POCUS scan at the bedside unlike many other studies that have used images from comprehensive echocardiography examinations obtained in echocardiography labs.^14–16^

Overall, this performance suggests that the device provides accurate and reliable measurements. The improved RMSEs compared with cardiologists may reflect the ability of the device to average across multiple beats and loops, with the cardiologists having been asked to quantify most measurements only once. This situation, however, mirrors the real-world in that cardiologists and other clinicians are often constrained by the need to manually obtain measurements while automated measurements can leverage a greater ability to average. A similar improvement in interobserver variability has previously been shown for conventional echocardiograms.^17^ The ability to average may also reflect the ability of the device to learn from the measurement styles of multiple cardiologists; in the context of electroencephalogram interpretation, this use of AI to average has been described as simulating a committee of independent experts.^27^

The one measurement where the device did not achieve the passing criteria was for RV fractional area change. For this measurement, however, the RMSE between the device and cardiologists was still better than between the cardiologists, reflecting the large variability that can be seen with this measurement and reinforcing the benefit that the device can provide through averaging. LV end diastolic diameter was the one measurement where the RMSE was greater between the device and cardiologists compared to between the cardiologists, although the RMSE between the device and cardiologists still achieved the secondary endpoint passing criterion.

POCUS provides clinicians with the ability to obtain critical diagnostic information at the bedside. For example, the knowledge of a patient’s LV ejection fraction or IVC maximal diameter can guide next management steps in a critical ill patient including whether to administer fluids.^28,29^ The Lumify (Philips) ultrasound probe, which was one of four assessed here, can connect directly to smart phones and tablets,^30^ while the Kosmos (EchoNous) ultrasound probe can connect directly to tablets.^31^ The use of the device with these ultrasound probes may be particularly beneficial in resource limited settings.

A key challenge with POCUS acquisition is that the image acquisition is often not as standardized as a conventional echocardiogram performed in an echocardiography laboratory. This study assessed how the device handles this challenge in multiple ways. Firstly, the device has quality control checks to ensure sufficient image quality and the presence of key points within images; the device did not provide a quantification for between 0.5% (for LV ejection fraction) and 13.5% (for LV end diastolic diameter) of cases for each measurement. Secondly, an image quality score was provided for each case; the RMSEs for image quality scores of ≤4 and >4 were similar for all measurements. Thirdly, the RMSEs were also similar for all measurements when the person acquiring the scan was a physician or a sonographer.

## Limitations

A key limitation of this study is that it was performed outside the clinical environment and therefore did not assess the impact of the device use on patient care and outcomes. Further to this point, the study did not assess the ability to generate the measurements in real-time with POCUS acquisition, where the device may provide the greatest clinical utility. Nevertheless, the images in this study were prospectively acquired at the bedside by mostly handheld devices and not as part of formal echocardiography studies obtained in an echocardiography lab; they therefore represented the real-world scenario where this device would be used. This study was also limited to four sites and the device will inevitably encounter new scenarios with clinical use, including further ultrasound probe manufacturers and models. For these reasons, the ongoing assessment of the device will be critical as it is deployed in the clinical environment.

## Conclusion

This AI-enabled device accurately calculated seven cardiac measurements on POCUS especially when benchmarked against inter-cardiologist variability. It could facilitate better clinical decision-making including in the acute care environment.

## Supporting information

STARD Checklist

## Data Availability

All data produced in the present study are available upon reasonable request to the authors.

## Acknowledgements

The authors thank the broader team members at the hospitals that acquired cases (Baruch Padeh Medical Center, Inova Health, Sheba Medical Center and Thomas Jefferson University Hospital), at Mass General Brigham AI and at AISAP LTD.

## Funding / Role of Funder

SFM, BCB, TS, MAH, ALM, INC, EL, ASS, VT, SMH, JRM, JW and JH were employees of Mass General Brigham and/or affiliated hospitals at the time of the study, which had received institutional funding from the sponsor (AISAP LTD) to conduct this project. PM and QZ were employees of Thomas Jefferson University and Inova Health System respectively, which had received institutional funding from the sponsor to conduct this project.

The sponsor was involved in the design and conduct of the study; preparation, review, or approval of the manuscript; and decision to submit the manuscript for publication. The sponsor was involved in the collection of POCUS cases, which were transferred to Mass General Brigham; the sponsor was otherwise not involved in the collection, management, analysis, and interpretation of the data.

## Conflicts of Interest

SMH received fees paid to her institution for core lab services from Cytokinetics and BMS; she is on the advisory board for Cytokinetics.

## Data Accountability

SFM had full access to all the data in the study and takes responsibility for the integrity of the data and the accuracy of the data analysis.

---

**Supplementary Table 1:** Views used to obtain each measurement. Abbreviations: A4, apical four chamber; PLAX, parasternal long axis.

| **Measurement** | **View** |
| --- | --- |
| Left ventricular ejection fraction | A4 / PLAX |
| Inferior vena cava maximal diameter | Subcostal |
| Left atrial area | A4 |
| Right atrial area | A4 |
| Left ventricular end diastolic diameter | PLAX |
| Right ventricular area (end of diastole and end of systole); to calculate right ventricular fractional area change | A4 |
| Aortic root diameter | PLAX |

**Supplementary Table 2:** Excluded cases. The cases were excluded when the cardiologists could not obtain the measurement or the device outputted “NULL” (see Methods for explanation). If a row exceeds a total of 200 cases, it indicates that a case was both excluded by the cardiologists and received a “NULL” device output. Abbreviations: IVC, inferior vena cava; LA, left atrial; LV, left ventricular; RA, right atrial; RV, right ventricular.

|  | Cardiologists<br>could not obtain | “NULL” device<br>output | Final cohort |
| --- | --- | --- | --- |
| LV ejection<br>fraction | 2 (1.0%) | 1 (0.5%) | 197 |
| IVC maximal<br>diameter | 16 (8.0%) | 25 (12.5%) | 175 |
| LA area | 3 (1.5%) | 10 (5.0%) | 188 |
| RA area | 5 (2.5%) | 18 (9.0%) | 178 |
| LV end diastolic<br>diameter | 1 (0.5%) | 27 (13.5%) | 173 |
| RV fractional area<br>change | 25 (12.5%) | 18 (9.0%) | 161 |
| Aortic root<br>diameter | 1 (0.5%) | 2 (1.0%) | 197 |

## Notes

### Funding Statement

This study was funded by AISAP LTD. SFM, BCB, TS, MAH, ALM, INC, EL, ASS, VT, SMH, JRM, JW and JH were employees of Mass General Brigham and/or affiliated hospitals at the time of the study, which had received institutional funding from the sponsor to conduct this project. PM and QZ were employees of Thomas Jefferson University and Inova Health System respectively, which had received institutional funding from the sponsor to conduct this project.

### Author Declarations

The acquisition of cases was approved by local Institutional Review Boards at each of Baruch Padeh Medical Center, Inova Health, Sheba Medical Center and Thomas Jefferson University Hospital. All patients provided informed consent. The subsequent diagnostic study involved transfer of de-identified cases to Mass General Brigham and was approved by the Mass General Brigham Institutional Review Board.

### Summary of Updates

This revision corrects an error in describing the training dataset.

